# Dynamics of SARS-CoV-2 with Waning Immunity in the UK Population

**DOI:** 10.1101/2020.07.24.20157982

**Authors:** Thomas Crellen, Li Pi, Emma L. Davis, Timothy M. Pollington, Tim C. D. Lucas, Diepreye Ayabina, Anna Borlase, Jaspreet Toor, Kiesha Prem, Graham F. Medley, Petra Klepac, T. Déirdre Hollingsworth

## Abstract

The dynamics of immunity are crucial to understanding the long-term patterns of the SARS-CoV-2 pandemic. Several cases of reinfection with SARS-CoV-2 have been documented 48–142 days after the initial infection and immunity to seasonal circulating coronaviruses is estimated to be shorter than one year. Using an age-structured, deterministic model, we explore potential immunity dynamics using contact data from the UK population. In the scenario where immunity to SARS-CoV-2 lasts an average of three months for non-hospitalised individuals, a year for hospitalised individuals, and the effective reproduction number after lockdown ends is 1.2 (our worst case scenario), we find that the secondary peak occurs in winter 2020 with a daily maximum of 387,000 infectious individuals and 125,000 daily new cases; three-fold greater than in a scenario with permanent immunity. Our models suggests that longitudinal serological surveys to determine if immunity in the population is waning will be most informative when sampling takes place from the end of the lockdown in June until autumn 2020. After this period, the proportion of the population with antibodies to SARS-CoV-2 is expected to increase due to the secondary wave. Overall, our analysis presents considerations for policy makers on the longer term dynamics of SARS-CoV-2 in the UK and suggests that strategies designed to achieve herd immunity may lead to repeated waves of infection as immunity to reinfection is not permanent.

## Introduction

### Public health context

As of the 1^st^ of July 2020, SARS-CoV-2 has infected at least 10 million people worldwide and resulted in over 500,000 deaths [1, 2]. Following the initial outbreak from a live animal market in Wuhan, China [3], the United Kingdom (UK) has been among the countries most severely affected; reporting over 310,000 cases and 44,000 deaths, which is among the highest per-capita rates [2, 4]. Since the 23^rd^ of March, nationwide non-pharmaceutical interventions (lockdown) have been in place to reduce social contacts by closing schools and shops; encouraging home working; and social distancing in public places. Similar measures have been in place in other European countries since late February 2020 with restrictions easing in France, Germany and Italy from May 2020. Within the European picture of disease control strategies, Sweden has been an outlier by placing fewer restrictions on social mixing while aiming to build up immunity in the population [5].

### Immune response to SARS-CoV-2

Following infection with the virus, hospitalised patients have an acute immune response where virus-specific IgM and IgG antibodies titres reach a maximum 15–21 and 22–27 days respectively after symptom onset [6, 7]. Antibodies raised in hospitalised patients and animal models against SARS-CoV-2 provide protection for at least several weeks following infection [8, 9], suggesting that immediate reinfection with the virus is unlikely. There is limited evidence that hospitalised patients with more severe symptoms show a greater antibody response [6, 9]. Asymptomatic individuals have a weaker IgG and specific antibody response to SARS-CoV-2 and are more likely to become seronegative following convalescence [10]. Antibody titres raised against related coronaviruses SARS-CoV and MERS-CoV have been shown to decay over time [11, 12]. Furthermore, immunity to seasonal circulating coronaviruses has been estimated to last for less than one year [13] and recovered individuals from coronavirus NL63 can become reinfected [14]. Concerns that immunity to SARS-CoV-2 may also wane therefore motivated the present study [15].

### Epidemiological modelling

Dynamic epidemiological models play a major role in shaping the timing and intensity of interventions against SARS-CoV-2 in the UK and elsewhere [16]. Many models or simulations have assumed that infected individuals recover with permanent immunity [16, 17, 18]. In such models the epidemic reaches extinction after running out of infected individuals, although they do not preclude a second wave of infections after lockdown [19]. If immunity wanes over a period of time, or recovered individuals have only partial immunity to reinfection, this substantially alters the dynamics of the system [20]. In the absence of stochastic extinction and demography (births and deaths) in a population with equal mixing where; *R*_0_ is the basic reproduction number; *γ* is the average duration of infection; and *ω* is the reciprocal of the average duration of immunity; the endemic equilibrium proportion of infected in the population *I*^*\**^, is given by (*R*_0_ − 1) *ω/γR*_0_ and thus, in the absence of interventions, the infection persists indefinitely when *R*_0_ *>* 1 [21].

In dynamic models which make the assumption of homogeneous mixing, the ‘classic’ threshold at which the proportion of the population with immunity prevents invasion by a pathogen is given by 1 − 1*/R*_0_ [21]. As *R*_0_ for SARS-CoV-2 is generally estimated between 2.4–5.6 [22, 23, 24], this equates to 58–82% of the population requiring immunity to eventually halt the epidemic. Serological studies conducted in affected countries to-date have reported the proportion of the population with antibodies against SARS-CoV-2 to be much lower than this figure [22, 25]. However, when more realistic non-homogeneous mixing is considered, the observed immunity threshold is lower than the classical threshold [26]. Recent studies have considered this question for SARS-CoV-2 [27, 28], with Britton *et al*. noting that the disease-induced population immunity threshold could be closer to 40% in an age-structured population when *R*_0_ is 2.5, rather than the 60% ‘classic’ population immunity threshold [28]. This phenomenon is driven by individuals that have more contacts, or greater susceptibility to the virus, getting infected earlier on and leaving the susceptible population; thus decelerating the growth of the epidemic.

### Models with waning immunity

Kissler *et al*. considered the dynamics of SARS-CoV-2 in the United States with seasonal forcing, homogeneous mixing and waning immunity that could be boosted by exposure to seasonal circulating betacoronaviruses [29]. Under these assumptions, the incidence of SARS-CoV-2 was predicted to rebound in winter months. Here we do not consider seasonality, but rather the dynamics of transmission in an age-structured population with different periods of waning immunity in the context of the UK emerging from lockdown.

We developed a discrete time gamma delay-distributed (susceptible-exposed-infectious-recovered-susceptible; SEIRS) model, which incorporates current knowledge about the natural history of the virus and the UK population. Our model accounts for symptomatic and asymptomatic transmission, and heterogeneity in both daily contacts and infection susceptibility by age group. We consider different durations of immunity for hospitalised patients (or those with more severe symptoms) compared to non-hospitalised patients (those with less severe symptoms). We use this model to explore a range of scenarios in the UK population in the context of stringent non-pharmaceutical interventions (lockdown) followed by more limited interventions over a two year period from March 2020, and the impact of immunity duration on the longer term disease equilibrium.

### Reinfection with SARS-CoV-2

After this analysis was presented to policymakers (see Accompanying Letter), the first case of reinfection with SARS-CoV-2 was reported in a 33 year old male in Hong Kong in August 2020; 142 days after the initial diagnosis [30]. Following the first infection in March, the patient was discharged from hospital after two negative PCR results. Genomic sequencing confirmed that the viruses in March and August were from distinct clades. Reinfections have since been reported in single cases in the USA [31], Ecuador [32] and Belgium [33] at 48, 63 and 93 days respectively from the first reported infections. Two asymptomatic reinfections were also reported in Indian healthcare workers 108 and 111 days from the initial infection [34]. The case of reinfection in the USA, in a 25 year old male from Nevada, was notable for the second infection causing more severe symptoms than the first, and led to the patient requiring supplemental oxygen and hospitalisation. While these reported cases confirm the biological plausibility of reinfection with SARS-CoV-2, the data are too sparse to infer an expected duration of immunity at the population level.

## Methods

### Model structure

We use current knowledge of the natural history of the virus to construct a plausible epidemiological model (Figure 1). We extend a previously published deterministic compartmental model which has provided general insights into the dynamics of the epidemic at a regional level for a range of scenarios [18]. The general framework of the model is given in Figure 1 and parameter values are shown in Table 1. The disease states for each age group are susceptible (*S*), exposed (*E*), symptomatic infectious (*I*^*S*^), asymptomatic infectious (*I*^*A*^), hospitalised recovered (*R*^*H*^), and non-hospitalised recovered (*R*^*N*^). Each state gives the number of individuals in the compartment at a given time *t*, with the sum over all the states at time *t* equal to the size of the UK population in 2018 (66.4 million).

**Table 1:** Summary of parameter values used in the modelled scenarios of SARS-CoV-2 transmission in the United Kingdom.

| Parameter name | Symbol | Estimate(s) | Details | Reference(s) |
| --- | --- | --- | --- | --- |
| Basic reproduction number | $R_0$ | 4.0 | - | [23] |
| Latency period mean | $\sigma$ | 4.5 days | - | [35, 55, 56] |
| Latency period shape | $m$ | 4 | - | [35, 55, 56] |
| Infectious period mean | $\gamma$ | 3.1 days | - | [35, 55, 56] |
| Infectious period shape | $n$ | 2 | - | [35, 55, 56] |
| Immune duration mean non-hospitalised | $\omega^N$ | $\infty, 365, 180, 90$ days | Varies by scenario | [11, 53, 57] |
| Immune duration mean hospitalised | $\omega^H$ | $\infty, 365$ days | Varies by scenario | [11, 53, 57] |
| Immune duration shape | $o$ | 2 | Centres distribution around mean | [11] |
| $\mathbb{P}(\text{asymptomatic} \mid \text{infection})$ | $\phi_i$ | $\leq 15$ yrs 0.75<br>$> 15$ yrs 0.5 | Varies by age group $i$ | [49] |
| $\mathbb{P}(\text{hospitalisation} \mid \text{infection})$ | $p_i$ | 0–0.26 | Varies by age group $i$ | [16, 44] |
| Effective reproduction number | $R_t$ | 1.2, 0.8 | During lockdown | [23, 58] |
|  |  | 0.9, 1, 1.1, 1.2 | After lockdown ends | Key assumption |
| Contact matrix | $C$ or $c_{ij}$ | Varies by age group | BBC survey | [47, 48] |
| Relative infectiousness of asymptomatic cases | $v$ | 0.5 | - | [16] |
| Relative age susceptibility | $\eta_i$ | $\leq 15$ yrs 0.4<br>$> 15$ yrs 1 | - | [52] |

**Figure 1:**
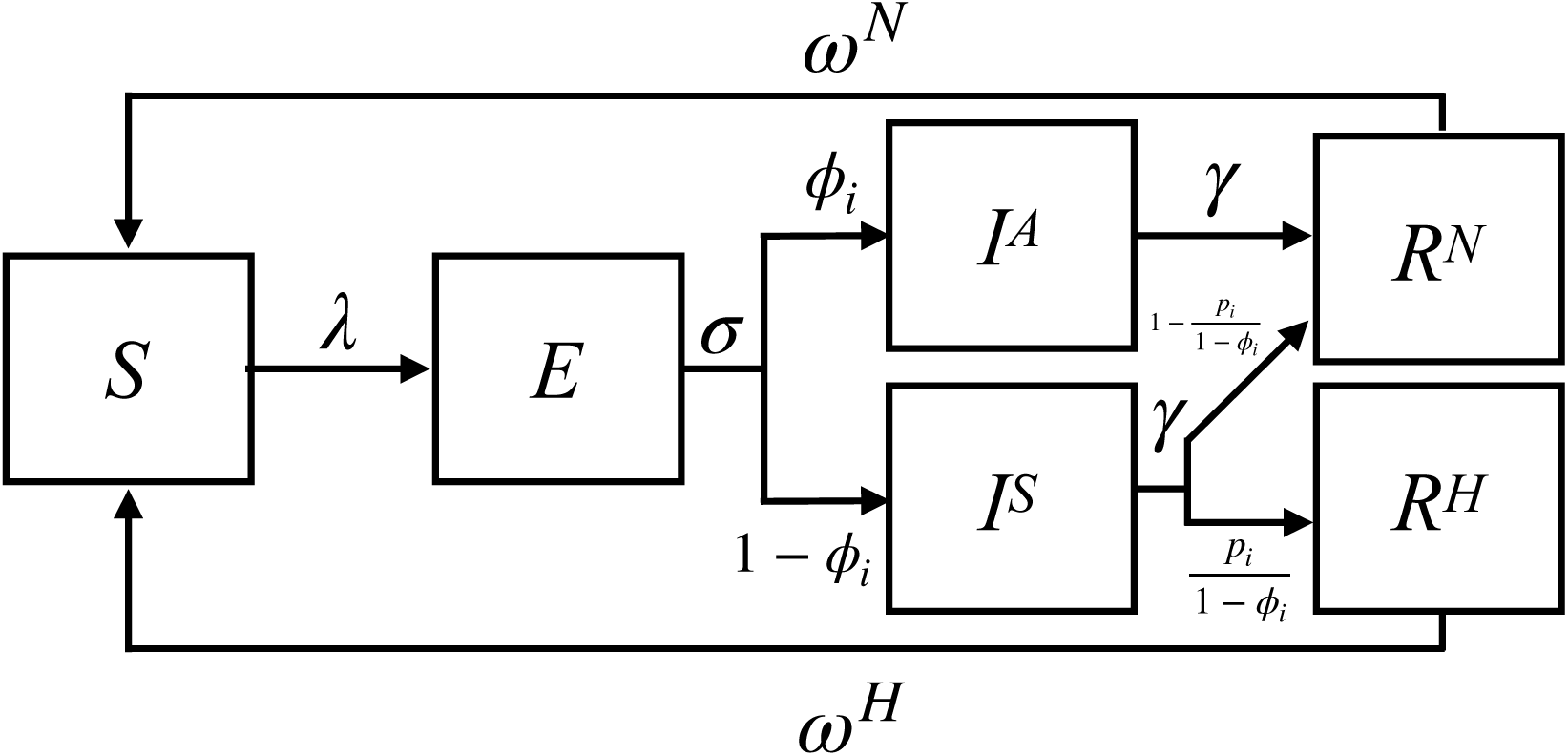
Flow diagram showing SARS-CoV-2 transmission model outline. Within age class *i*, the disease states are susceptible (*S*), exposed (*E*), symptomatic infectious (*I*^*S*^), asymptomatic infectious (*I*^*A*^), hospitalised recovered (*R*^*H*^), and non-hospitalised recovered (*R*^*N*^). The rate parameters *λ* represent the force of infection acting on an individual in age class *i* (Equation 8), *σ* represents the mean latency period, *γ* the mean infectious period, *ω*^*N*^ the mean duration of immunity for non-hospitalised recovered individuals, and *ω*^*H*^ the mean duration of immunity for hospitalised recovered individuals. The probabilities *φ*_*i*_ give the proportion of individuals in age group *i* that are asymptomatic following infection and *p*_*i*_ the proportion in age group *i* that require hospitalisation (or have severe symptoms) following infection. All model parameters are given in Table 1 and state transitions are shown in Equations 1 to 6.

### Distributed natural history of infection

The mean latent and infectious periods for SARS-CoV-2 have been estimated as 4.5 days and 3.1 days respectively, using viral load data and the timing of known index and secondary case contacts (Figure 2) [35]. The probability mass of the latent and infectious period distributions are centred around the mean, therefore we consider that gamma distributions with an integer shape parameter, also known as Erlang distributions, give more realistic waiting times than exponential distributions which have a mode of zero [36, 37, 38]. We decided to use a discrete time implementation (difference equations), as this permits greater flexibility in implementing interventions compared to differential equations. Using difference equations results in a discretised approximation of the Erlang distribution compared with continuous time models, with the output converging as the time step approaches zero [39, 40]. We used a time step of 0.01 days to ensure that the infection dynamics closely approximate a continuous time model.

**Figure 2:**
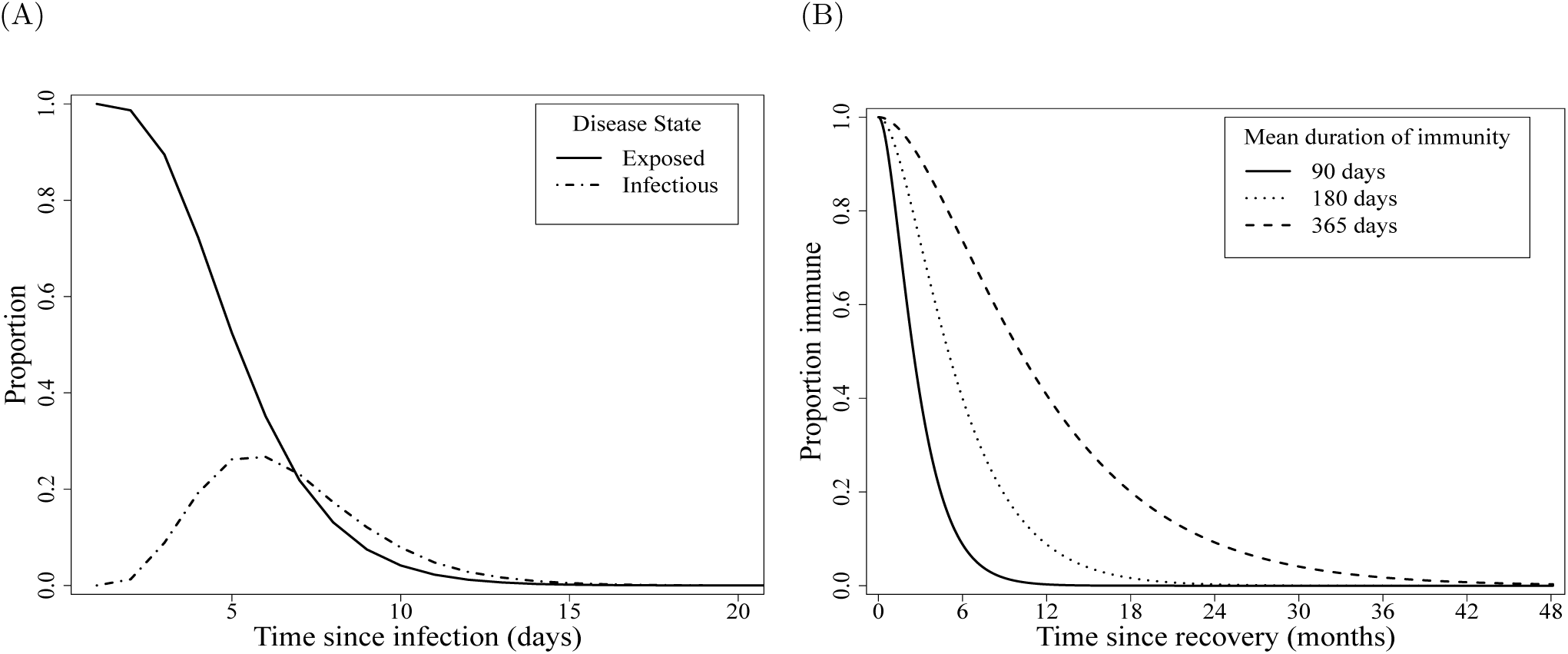
Probabilities for time spent in each state given Erlang (gamma) distributed waiting times. A. Proportion of individuals in exposed and infectious classes since time from infection. Time exposed and time infectious have mean durations of 4.5 and 3.1 days respectively. B. Proportion of individuals immune since recovery, where time immune has mean durations of 90, 180 or 365 days depending on the scenario.

### Transmissibility and infectivity

Estimates of the transmissibility of the virus in the UK at the beginning of the epidemic have ranged from 2.4–5.6 [23, 41, 42], therefore we assume that *R*_0_ at the beginning of the epidemic in the UK population is 4.0 [23]. Non-pharmaceutical interventions have been shown to bring the effective reproduction number (*R*_*t*_) below one, and in some settings have led to local elimination of the virus [22, 23].

Testing performed in closed populations suggests that 40-50% of SARS-CoV-2 infections may be asymptomatic [43, 44, 45], while data from contact tracing shows transmission can occur from asymptomatic individuals [46]. We make the assumption that asymptomatic individuals (*I*^*A*^) have 0.5 the infectiousness of symptomatic individuals (*I*^*S*^) [6, 16].

The UK population shows variable contact rates by age [47, 48] and, while studies show mixed results, evidence is accruing that children have a lower susceptibility to acquiring the infection than adults [49, 50, 51]. We assume that children (*≤*15 years) have 0.4 times the susceptibility of adults [52].

### Immunity following infection

We allow the duration of immunity to differ for recovered individuals with severe symptoms that are hospitalised (*R*^*H*^) versus those with less severe symptoms that are not hospitalised (*R*^*N*^), as there is evidence from SARS-CoV-2 and other coronaviruses that individuals with milder symptoms may have a lower antibody response [53]. Among the reported cases of reinfections, all individuals showed at most ‘mild’ symptoms during the first infection [54], supporting the notion that infections which cause more severe symptoms could generate longer lasting immunity.

Epidemic transitions for age group *i* at time *t* + 1 are given by:

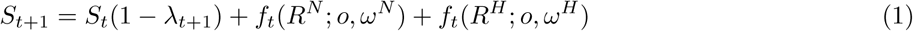

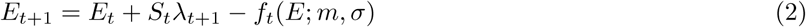

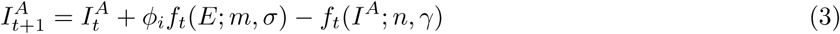

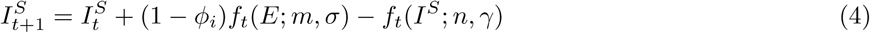

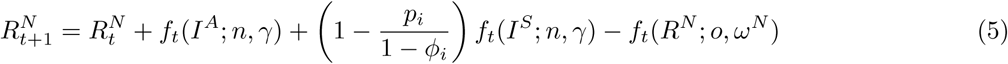

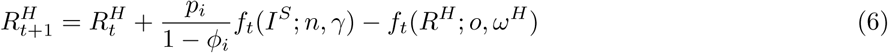

The function notation *f*_*t*_(*X*; *k, µ*) represents the output of the discretised Erlang delay distribution from states *E, I*^*S*^, *I*^*A*^, *R*^*H*^ and *R*^*N*^. Our model code contains *k* sub-compartments for each Erlang distributed disease state, with a rate *k/µ* between sub-compartments, and an overall waiting time of *µ*. The function notation is used in equations 1–6 for shorthand convenience instead of writing out *k* difference equations for each disease state. While we do not calculate probability distributions within the model, as a simplified example; if *n* individuals enter state *X* at a single time point *t*, by time *t* + *τ* there will be remaining *n*(1 − *g*(*τ, k, µ*)); where *g*(*τ, k, µ*) gives the cumulative Erlang distribution with shape parameter *k* and mean duration *µ* [39].

The next generation matrix (*K* = *k*_*ij*_) gives the expected number of secondary infections in age group *i* resulting from contact with an index case in age group *j*:

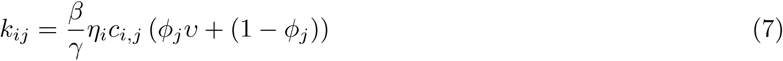

where *η*_*i*_ is the relative susceptibility of age group *i*; *υ* the relative infectiousness of asymptomatic cases; and *c*_*i,j*_ is the average number of daily contacts between a single individual in age group *j* and all individuals in age group *i* [48] (Table 1). The basic reproduction number (*R*_0_) is given by the spectral radius *ρ*(*K*) which is the largest absolute eigenvalue of *K*. As we specify the value of *R*_0_, the transmission parameter *β* is left as a free parameter which is scaled to the correct value. The force of infection acting on a single individual in age group *i* at time *t* + 1 (*λ*_*i,t*+1_) is given by:

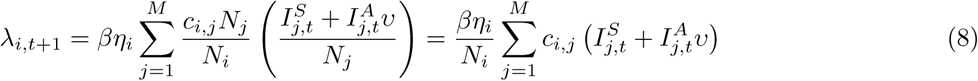

where *M* is the number of discrete age groups (*M* = 15); and *N*_*i*_ gives the population size of age group *i*.

### Immunological scenarios

Using data and timing of events from the UK epidemic, we explore four scenarios with varying average durations of immunity to SARS-CoV-2 (Figure 2).

S1. **Permanent:** Where immunity is lifelong for both hospitalised (*R*^*H*^) and non-hospitalised (*R*^*N*^) cases.

S2. **Waning (12 months):** Where immunity is lifelong for hospitalised cases and has an average duration of 365 days for non-hospitalised cases.

S3. **Waning (6 months):** Where immunity is lifelong for hospitalised cases and has an average duration of 180 days for non-hospitalised cases.

S4. **Short-lived:** Where immunity lasts, on average, 365 days for hospitalised cases and 90 days for non-hospitalised cases.

### UK-specific parameterisation

All scenarios are initialised with 2968 infected individuals in early March 2020 [59], which is when the majority of importation events into the UK are likely to have occurred [59]. Intervention measures are initiated on the 23^rd^ of March (date the UK nationwide lockdown started), with an immediate reduction in the effective reproduction number (expected number of secondary cases from an index case at time *t*; *R*_*t*_) to 1.2 for a three week period, followed by a further reduction in *R*_*t*_ to 0.8 until lockdown measures are eased on 15^th^ June [58]. After this time, *R*_*t*_ is brought to 0.9, 1, 1.1 or 1.2 until March 2022. We considered the majority of our analysis over a, relatively short, two year period to explore the epidemic up to a secondary peak; beyond this point the dynamics are likely to be altered depending on further interventions or changes to *R*_*t*_. As we simulate disease dynamics over a relatively short period of time, we do not consider demography (births and deaths) or transitions between age classes (ageing). To obtain equilibrium values, we simulated epidemic trajectories for up to five years.

The UK contact matrix (average daily contacts between a single individual in age group *j* with individuals in age group *i*) comes from a ‘citizen science’ project for the BBC, in which individuals in the UK population provided detailed information on their daily contacts in the home, in the workplace, at school and in other settings [47, 48]. The contact matrix is altered to account for changes to contact patterns during and after the main intervention period [56]. During the lockdown, home; work; school; and other contacts are reduced to 0.8, 0.3, 0.1 and 0.2 respectively of their baseline values. This reflects the school closures for all children, except for those of key workers, and that workers were encouraged to work from home. Reduction in home contacts accounts for the absence of visitors to the home during the lockdown. In the post-lockdown phase, home; work; school; and other contacts are scaled to 1, 0.8, 0.85 and 0.75, respectively, of their baseline values to reflect limited social distancing measures that are likely to be in place until at least the end of 2021.

Analysis was performed in R version 4.0.2. We present figures from model output in the text to the nearest thousand. Code to reproduce the analysis with user-specified parameters is available at https://github.com/tc13/covid-19-immunity.

## Results

### Heterogeneity in transmission

The epidemic is driven by the rate of infectious contacts between individuals in different age groups. This is described by the next generation matrix in which the average number of secondary cases generated by an index case in age group *j* is the summation of row *j* (Equation 8 & Figure 3). At the beginning of the epidemic, when SARS-CoV-2 is spreading rapidly, all age groups are involved in transmission; in particular those aged 20–39 years. An index case in the 20–24 age group, for instance, is expected to generate an average of 4.3 secondary cases at baseline. As lockdown measures come into force this dramatically reduces the expected number of secondary cases due to fewer contacts and a lower probability of infection given contact. The average number of secondary cases from an individual aged 20–24 during lockdown drops to 0.9 and the transmission parameter *β*, which captures the probability of infection given contact, is decreased from 0.18 at baseline to 0.11. In the post-lockdown period daily contacts are increased to a higher proportion of their baseline values (see Methods); in order to keep the reproduction number equal to the dominant eigenvalue of the next generation matrix, *β* is consequently reduced to 0.05 when *R*_*t*_ =0.9 and to 0.07 when *R*_*t*_ =1.2. This implies that, to maintain *R*_*t*_ below one when more contacts are occurring in the population post-lockdown, the probability that contact results in infection will need to be reduced.

**Figure 3:**
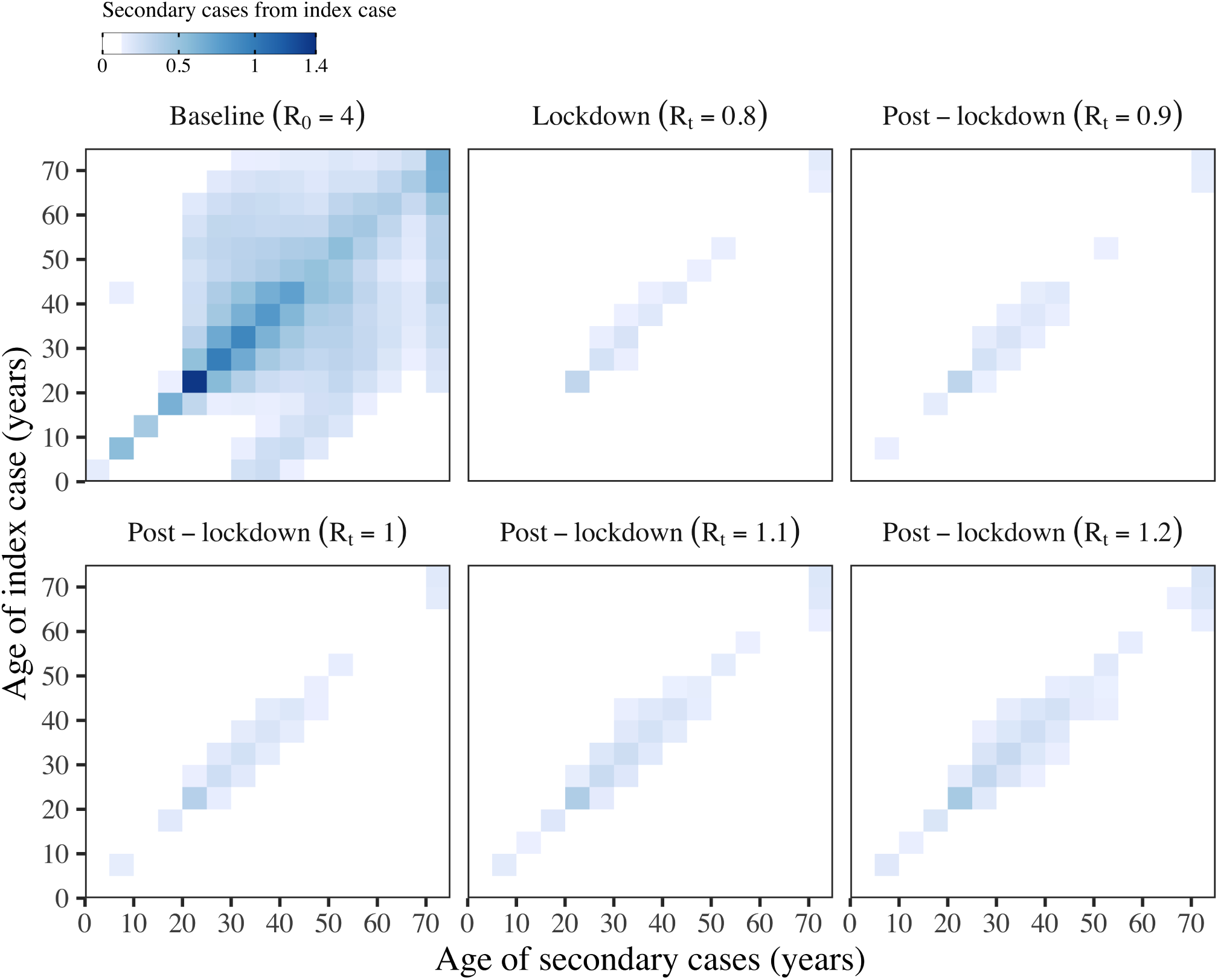
Next generation matrix (*K* = *k*_*ij*_) showing the number of secondary cases generated by an index case from age group *j* (rows) in age group *i* (columns). The matrices are shown for different time points; at baseline before the implementation of interventions; during the lockdown period; and in the post-lockdown period when the effective reproduction number (*R*_*t*_) rises to 0.9–1.2. The average number of secondary cases generated by an index case from age group *j* is the summation of row *j*.

### Infection dynamics

For the first 100 days until the end of the lockdown, the infection dynamics are equivalent across the four immunity scenarios S1–S4 (Figure 4, panels A, C, E & G). After this time the dynamics depend on both the rate at which recovered individuals lose immunity and become susceptible again, and the post-lockdown *R*_*t*_.

**Figure 4:**
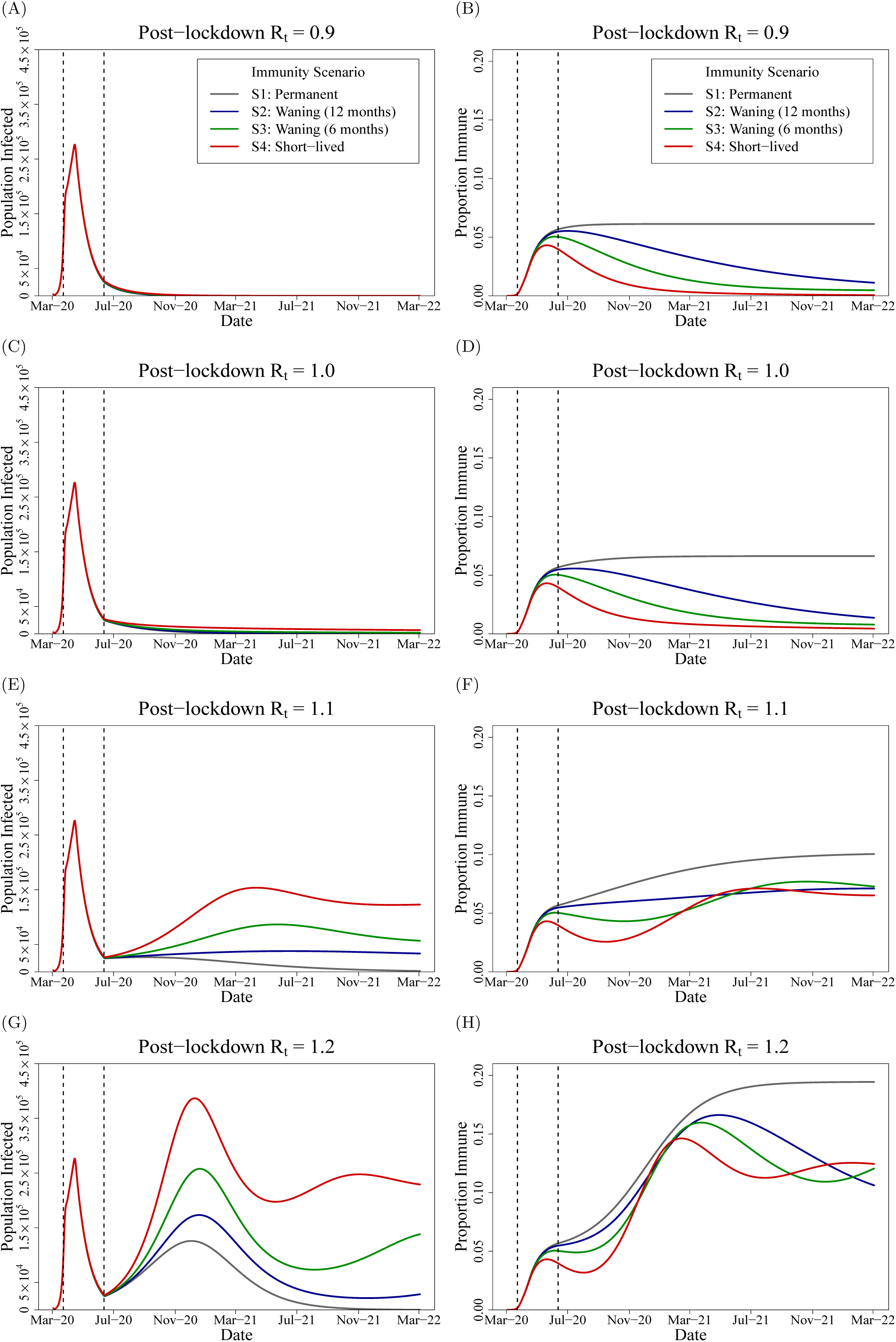
Projections from immunity scenarios S1–4 with post-lockdown *R*_*t*_ ranging from 0.9–1.2. Left panels show the number of infected, both asymptomatic and symptomatic (*I*^*A*^ + *I*^*S*^), with SARS-CoV-2 in the UK population over time. Right panels show the proportion of the UK population with immunity (compartments *R*^*H*^ + *R*^*N*^). Dashed vertical lines indicate the lockdown period; 23^rd^ of March–15^th^ of June 2020.

Given our model and parameters, on the first day the intervention is imposed (23^rd^ March 2020) there are 108,000 new SARS-CoV-2 cases, which is within the 95% credible interval (CrI) of new cases estimated for the UK on that day (95% CrI 54,000–155,000 [60]), and 75,000 people are infectious (infected compartments *I*^*A*^ + *I*^*S*^) on this date. From the 3^rd^ of March until the 23^rd^ of March there are 559,000 cumulative cases across all age groups and 524,000 in adults ≥19 years, which is within the credible interval for an estimate of cumulative cases in this period (95% CrI 266,000–628,000 [60]). When most of the lockdown measures were eased in June, 5.7% of the total population and 6.8% of adults aged ≥19 years have immunity to SARS-CoV-2 (in recovered classes *R*^*H*^ and *R*^*N*^), which is comparable to estimates of antibody levels in the UK population, estimated as 6.8% of blood donors on 24^th^ May 2020 (95% confidence interval 5.2–8.6%; individuals ≥18 years [25]).

### Secondary peak in infections

A secondary peak in infections is expected in spring 2021 where *R*_*t*_ = 1.1 or winter 2020 where *R*_*t*_ = 1.2 (Figure 4, panels E & G). The height of the secondary peak is determined by the rate at which immunity is lost. In our worst case scenario (S4: short-lived immunity) where immunity lasts an average of three months for non-hospitalised patients, a year for hospitalised patients and *R*_*t*_ following lockdown is 1.2, then the secondary peak will exceed the initial peak with a maximum of 387,000 infectious individuals and 125,000 daily new cases in December 2020. This is three-fold greater than the number of new cases in scenario S1 where immunity is permanent; the maximum number of infectious individuals in the secondary peak is 126,000 and there are 41,000 daily new cases (Figure 4G). We note that the timing of the secondary peak in infection curves across immunological scenarios are closely synchronised and in winter 2020–21. This synchrony and timing is also observed during the epidemic when values of *R*_*t*_ post-lockdown are greater than 1.2 (explored for values of *R*_*t*_ from 1.3 to 2.0).

When *R*_*t*_ following lockdown is 1.1 the differences between the immunity scenarios is even more pronounced with a ten-fold difference in the height of the secondary peak between a scenario of permanent immunity and one of short-lived immunity, however the secondary peak does not exceed the height (maximum number of infectious individuals) of the initial wave in any scenario. When immunity wanes rapidly, the second peak is observed in April 2021 with a maximum of 154,000 infectious individuals and 50,000 daily new cases. By contrast when immunity is permanent, the number of new infections decays slowly rather than accelerates and there are projected to be a maximum of only 15,000 infectious individuals and 5,000 daily new cases in April 2021 (Figure 4E).

### Population immunity

The dynamics of population immunity (recovered compartments *R*^*H*^ +*R*^*N*^) are similarly shaped by the expected duration of immunity against SARS-CoV-2 and the post-lockdown *R*_*t*_.

Immunity decays from midway through the lockdown period in scenarios S2–S4 of waning (12 or 6 months) and short-lived immunity and resurges following a secondary wave of infection if *R*_*t*_ *>* 1 (Figure 4, panels F & H). After lockdown, a fall in the proportion of the population immune to the virus is observed until autumn 2020 for all values of *R*_*t*_, after which point the secondary wave, if *R*_*t*_ *>* 1, causes the proportion of the population immune to rise again. This suggests that longitudinal serological surveys to detect waning immunity would be most informative when conducted in the period June–October 2020.

### Consequences of age structure

The large differences in the heights of the secondary peaks when *R*_*t*_ *>* 1 between immunological scenarios (Figure 4, panels A, C, E & G) can be explained by the heterogeneity in transmission (see the next generation matrix in Figure 3). Infectious and immune cases as a proportion of the total age group are shown in Figure 5 for scenarios S1 & S4 of permanent and short-lived immunity where *R*_*t*_ = 1.2 following lockdown. A higher proportion of individuals aged between 20–39 are infected early in the epidemic, and this leads to 12.4–13.9% of individuals in these age groups having antibodies by October 2020 whe’n immunity is life-long (Figure 5B). When immunity wanes, however, by October 2020 this drops to 6.7–7.8% (Figure 5D), thus replenishing the pool of susceptible individuals with the age groups that disproportionately drive transmission. This causes the secondary peak of infectious cases to rise more rapidly and to a greater height when immunity wanes rapidly (Figure 5C), compared with permanent immunity (Figure 5A). Our models suggest that the age distribution of cases in the epidemic will not change greatly over time; as seen in Figure 5 the ordering of the proportion of each age group infected remains constant in both scenarios of permanent and short-lived immunity.

**Figure 5:**
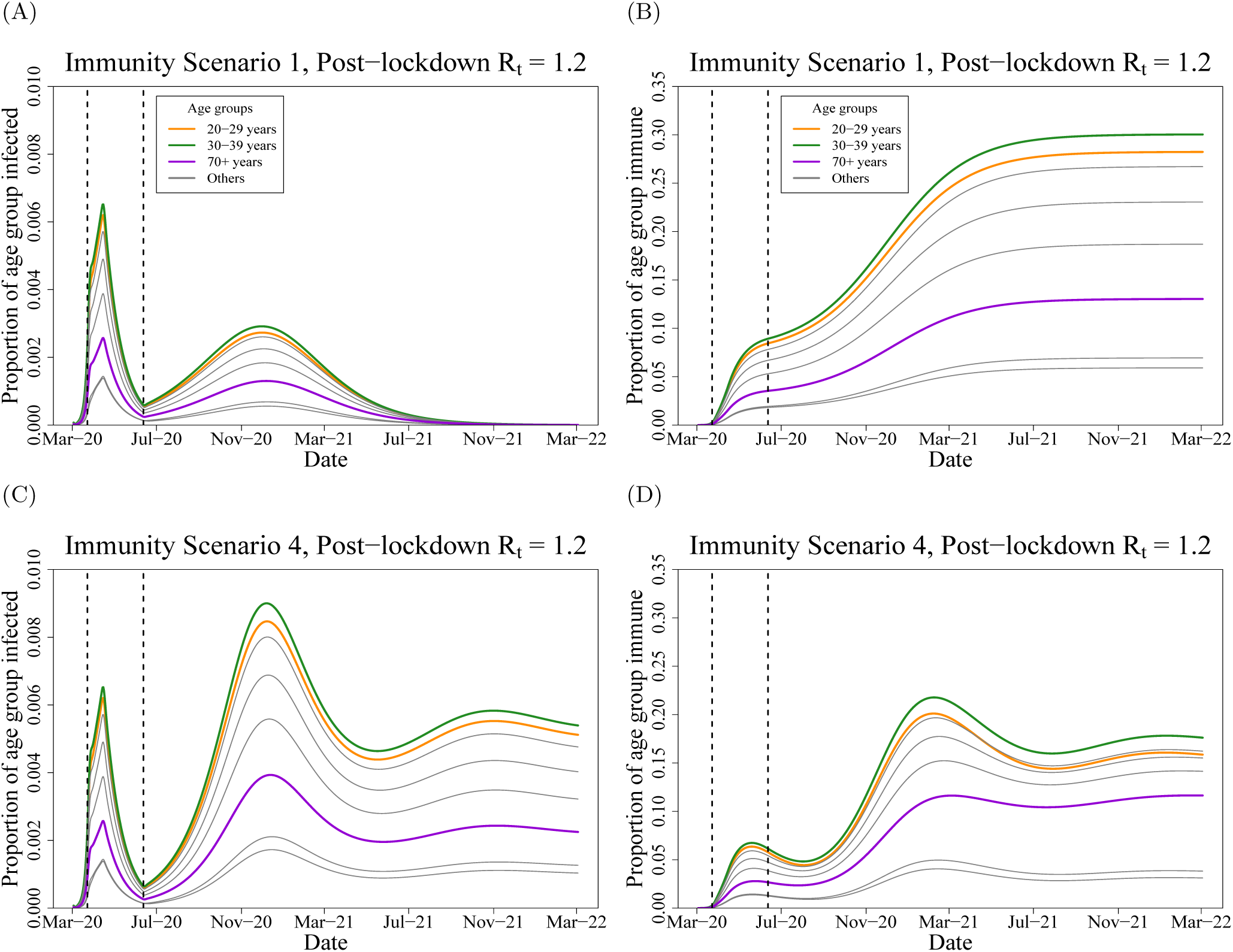
Projections from immunity scenarios S1 & S4 with post-lockdown *R*_*t*_ of SARS-CoV-2 at 1.2 in the UK population over time. Left panels show the proportion of each age group infected, for both asymptomatic and symptomatic (*I*^*A*^ + *I*^*S*^) individuals. Right panels show the proportion of the each age group with immunity (compartments *R*^*H*^ + *R*^*N*^). Dashed vertical lines indicate the intervention (lockdown) period; 23^rd^ of March – 15^th^ of June 2020.

### Longer term dynamics: extinction or endemic equilibrium

We explored the impact of waning immunity and *R*_*t*_ on the equilibrium values for the different simulations over a longer, five year, period until March 2025 (Table 2). If the post-lockdown *R*_*t*_ is suppressed below one following lockdown, then the differences in immunity will have less impact on the longer-term infection dynamics, assuming no imported cases, as transmission of SARS-CoV-2 becomes unsustainable and the virus reaches extinction between April–November 2021 depending on the immunity scenario. In simulations where *R*_*t*_ equals one, if immunity is permanent then the epidemic becomes extinct in May 2022. When immunity wanes there is no secondary peak (Figure 4C), however the infections persist at a low level of endemicity equivalent to 107, 236 and 1,122 daily cases in immunity scenarios S2–S4, respectively. For larger values of *R*_*t*_, and where immunity wanes, the system oscillates with subsequent peaks of infection over the next five years until a steady state is reached. We find that, if *R*_*t*_ = 1.2 post-lockdown and immunity is short-lived, there is the potential for over 73,000 new cases daily; 5,000 hospitalisations; and 913 intensive care unit (ICU) admissions (calculated as 17% of all hospitalised cases [61]) at endemic equilibrium (January 2025), which would be sufficient to overwhelm contact tracing services and ICU capacity [62, 63].

**Table 2:** Values at equilibrium from the modelled scenarios for SARS-CoV-2 in the United Kingdom, explored over a five year horizon (March 2020 to March 2025). ^1^Effective reproduction number of SARS-CoV-2 after lockdown. ^2^Assumed duration of immunity for hospitalised and non-hospitalised individuals, see Methods for details of immunity scenarios S1–S4. ^3^Number of individuals newly infected with SARS-CoV-2 that enter the exposed *E* state. ^4^Number of symptomatic individuals with SARS-CoV-2 that enter the recovered hospitalised (*R*^*H*^) state. ^5^Number of hospitalised individuals admitted to intensive care units (ICU), under the assumption that 17% of hospitalised cases in the UK require care in high dependency units [61]. ^6^Either when the number of daily new cases drops below one (extinction), or when the daily new cases are the same integer value over a sustained period (endemic equilibrium). If models take longer than five years to reach a steady state, the values on the 1^st^ of March 2025 are reported.

| $R_t$<sup>1</sup> | Immunity scenario<sup>2</sup> | Daily cases<sup>3</sup> | Daily hospitalisations<sup>4</sup> | Daily ICU admissions<sup>5</sup> | Date equilibrium reached<sup>6</sup> |
| --- | --- | --- | --- | --- | --- |
| 0.9 | S1: Permanent | 0 | 0 | 0 | April 2021 |
|  | S2: Waning (12 months) | 0 | 0 | 0 | June 2020 |
|  | S3: Waning (6 months) | 0 | 0 | 0 | September 2021 |
|  | S4: Short-lived | 0 | 0 | 0 | November 2021 |
| 1.0 | S1: Permanent | 0 | 0 | 0 | April 2022 |
|  | S2: Waning (12 months) | 71 | 5 | 1 | October 2022 |
|  | S3: Waning (6 months) | 217 | 16 | 3 | After 1<sup>st</sup> March 2025 |
|  | S4: Short-lived | 1,160 | 84 | 14 | After 1<sup>st</sup> March 2025 |
| 1.1 | S1: Permanent | 0 | 0 | 0 | June 2024 |
|  | S2: Waning (12 months) | 9,117 | 654 | 111 | After 1<sup>st</sup> March 2025 |
|  | S3: Waning (6 months) | 14,867 | 1,047 | 178 | After 1<sup>st</sup> March 2025 |
|  | S4: Short-lived | 39,945 | 2,887 | 491 | May 2023 |
| 1.2 | S1: Permanent | 0 | 0 | 0 | November 2022 |
|  | S2: Waning (12 months) | 19,826 | 1,443 | 245 | After 1<sup>st</sup> March 2025 |
|  | S3: Waning (6 months) | 27,488 | 1,898 | 323 | After 1<sup>st</sup> March 2025 |
|  | S4: Short-lived | 73,977 | 5,370 | 913 | March 2024 |

## Discussion

Despite only 6.8% of the adult UK population having immunity against SARS-CoV-2 in our simulation at the end of the lockdown, the modelled scenarios suggest that, if this acquired immunity wanes over time, there are substantive differences to the subsequent infection dynamics. Waning immunity impacts on the height of the secondary peak and, in the absence of future interventions, establishes the virus at levels of endemic equilibrium that could overwhelm contact tracing services and ICU capacity [62, 63].

We predict that surveys to detect waning immunity at the population level would be most effective when carried out in the period between the end of lockdown and autumn 2020, as after this point an upsurge in cases is expected that will increase the proportion of the population with immunity to SARS-CoV-2. In particular, this will allow evaluation of whether specific antibodies generated against the virus are short-lived if reductions in antibody prevalence are observed at the population level.

We find that transmission is driven disproportionately by individuals of working age, and subsequently a higher proportion of individuals aged 20–39 years become infected early in the pandemic and subsequently develop immunity (Figures 3 & 5). This prediction is borne out by serological data from Switzerland, which showed that individuals aged 20–49 years were significantly more likely to be seropositive in May 2020 compared with younger and older age groups [64]. We postulate that ‘key workers’ in the UK population who have continued to work during the lockdown are more likely to have antibodies against SARS-CoV-2. Higher immunity among individuals of working age has the effect of slowing the subsequent epidemic when immunity is permanent. Conversely, when immunity wanes, previously infected individuals of working age rejoin the susceptible pool and so contribute again to transmission; leading to a high growth rate and a larger secondary peak of infected cases. In these circumstances, efforts to suppress transmission will be challenging in the absence of a transmission-blocking vaccine [15]. We note that the model structure developed here is capable of simulating the impact of vaccination with a vaccine that provides temporary transmission-blocking immunity, and could be used to predict the optimal timing for booster shots.

The projected trajectory of the epidemic after lockdown is highly sensitive to the effective reproduction number, with model behaviour for values of *R*_*t*_ slightly above or below one displaying qualitatively different dynamics (Figure 4). Although we do not describe model output for higher values of *R*_*t*_, the dynamics over a wider range of effective reproduction numbers post-lockdown (1.3–2.0) are qualitatively the same as when *R*_*t*_ =1.2. This shows the importance of timely and accurate estimates of *R*_*t*_ to inform control strategies, and ensuring widespread community testing and contact tracing is in place. Our calculations show that to suppress *R*_*t*_ below one when contact rates rise to a higher fraction of baseline (pre-lockdown) values, the probability of infection given contact (represented by the *β* parameter), must drop by around half. Interventions that have the potential to reduce the probability of infection include social distancing; regular hand washing; and the wearing of face masks outside the home [65].

Our study reinforces the importance of better understanding SARS-CoV-2 immunity among recovered individuals of different ages and disease severity. In scenarios where immunity wanes and *R*_*t*_ following lockdown is greater than one, the SARS-CoV-2 epidemic never reaches extinction due to herd immunity, but rather the number of infected cases oscillates with subsequent waves of infection before reaching endemic equilibrium (Table 2). Even in simulations where the post-lockdown reproduction number only narrowly exceeds one (*R*_*t*_ =1.1), if immunity wanes over an average of one year for severe cases and three months for non-severe cases, this is projected to lead to an equilibrium state of nearly 40,000 daily new cases and 500 daily admissions to intensive care. Policy strategies aiming to achieve herd immunity are therefore risky [5]. As individuals can lose their immunity and become reinfected with SARS-CoV-2, a stable population immunity threshold can never be reached in the absence of a vaccine with lifelong efficacy [21]. The establishment of an endemic equilibrium state is dependent on no future interventions or changes to *R*_*t*_, which we consider unlikely as policy makers and public health agencies are likely to react to future outbreaks with localised control measures.

One of the strengths of our study is that the model is calibrated to key features of the UK epidemic. While we did not explicitly fit to data, new cases at the start of the lockdown; cumulative cases in March; and the proportion of the adult population with antibodies to SARS-CoV-2 are highly comparable between our output and current estimates [25, 60]. We used contact matrices from a comprehensive study of contact patterns in the UK population [47] in addition to demographic data from the Office for National Statistics, to give our simulations the best chance of capturing realistic age-specific transmission patterns in the UK population.

Plausible estimates on which to base expectations for the duration of immunity are sparse in the current literature. Rosado *et al*. estimated that antibodies could wane in 50% of recovered individuals after one year [57], which is similar to the estimated duration of immunity against seasonal circulating coronaviruses [13, 29].

Even with this consideration, there are many probability distributions that can be used to capture a median duration of immunity, and our selection of an Erlang distribution with a shape parameter of two is somewhat arbitrary. Our assumptions on the duration of the latent and infectious periods are more closely informed by estimates from data [35, 55, 56]. We made the decision to capture the expected duration of these states as a discretised Erlang distribution rather than the, more conventional, exponential distribution. This has the benefit of closely replicating fitted gamma or log-normal distributions within a compartmental model [36], and has important implications for the dynamics of the epidemic [66, 67]. We make a number of assumptions regarding the natural history of the virus, such as the relative susceptibility of children compared with adults and the relative infectiousness of symptomatic versus asymptomatic cases based on the current literature [52, 49]. Future empirical studies are likely to add to and further refine these epidemiological parameters. After we completed the analysis, a study of 37 asymptomatic individuals in China were found to have a longer period of viral shedding when compared with symptomatic individuals [10]. While viral shedding is not necessarily indicative of transmission potential [7], if these findings are replicated in larger studies this may suggest a need to use different durations of infectiousness for asymptomatic and symptomatic infections in subsequent models.

We have aimed to capture future infection dynamics at a national level in the UK under a range of scenarios. Our analysis is limited by not considering regional differences in transmission rates, for instance through a patch (metapopulation) model [48], or a stochastic approach that allows for local extinction events [21]. There are no deaths in our model, either from demography or infection. Accounting for mortality would mainly affect dynamics in the oldest age group (over 70 years) [16, 61], and the higher probability of disease-induced mortality would reduce the build up of immunity in this age group (Figure 5D). We also do not explicitly consider transmission in settings such as schools, hospitals or care homes, although such dynamics may be captured indirectly through the contact matrix. Given the simplicity of the model structure, we advise against treating the output as an exact prediction of the future. In addition to the limitations listed above, the epidemic trajectory will be substantially altered by any future interventions such as a return to full lockdown conditions, or intensive contact tracing and isolation [29, 68]. We emphasise that the average duration of immunity following recovery impacts substantially on epidemic growth following the easing of non-pharmaceutical interventions, and that slight differences in *R*_*t*_ above or below one can lead to qualitatively different infection dynamics.

## Data Availability

This is a modelling study and does not use primary data. Open access code to reproduce the analysis is provided.

https://github.com/tc13/covid-19-immunity

## Acknowledgements

This work was undertaken as a contribution to the Rapid Assistance in Modelling the Pandemic (RAMP) initiative, coordinated by The Royal Society. The authors thank Prof. Julia Gog and Dr. Rosalind Eggo for their helpful comments and input.

## Open access

Anyone can share and adapt this article provided they give credit and link to this article’s DOI and CC BY 4.0 licence (creativecommons.org/licences/by/4.0).

